# Comorbidity and its impact on 1,590 patients with COVID-19 in China: A Nationwide Analysis

**DOI:** 10.1101/2020.02.25.20027664

**Authors:** Wei-jie Guan, Wen-hua Liang, Yi Zhao, Heng-rui Liang, Zi-sheng Chen, Yi-min Li, Xiao-qing Liu, Ru-chong Chen, Chun-li Tang, Tao Wang, Chun-quan Ou, Li Li, Ping-yan Chen, Ling Sang, Wei Wang, Jian-fu Li, Cai-chen Li, Li-min Ou, Bo Cheng, Shan Xiong, Zheng-yi Ni, Jie Xiang, Yu Hu, Lei Liu, Hong Shan, Chun-liang Lei, Yi-xiang Peng, Li Wei, Yong Liu, Ya-hua Hu, Peng Peng, Jian-ming Wang, Ji-yang Liu, Zhong Chen, Gang Li, Zhi-jian Zheng, Shao-qin Qiu, Jie Luo, Chang-jiang Ye, Shao-yong Zhu, Lin-ling Cheng, Feng Ye, Shi-yue Li, Jin-ping Zheng, Nuo-fu Zhang, Nan-shan Zhong, Jian-xing He, on behalf of China Medical Treatment Expert Group for COVID-19

## Abstract

**Objective:** To evaluate the spectrum of comorbidities and its impact on the clinical outcome in patients with coronavirus disease 2019 (COVID-19).

**Design:** Retrospective case studies

**Setting:** 575 hospitals in 31 province/autonomous regions/provincial municipalities across China

**Participants:** 1,590 laboratory-confirmed hospitalized patients. Data were collected from November 21^st^, 2019 to January 31^st^, 2020.

**Main outcomes and measures:** Epidemiological and clinical variables (in particular, comorbidities) were extracted from medical charts. The disease severity was categorized based on the *American Thoracic Society* guidelines for community-acquired pneumonia. The primary endpoint was the composite endpoints, which consisted of the admission to intensive care unit (ICU), or invasive ventilation, or death. The risk of reaching to the composite endpoints was compared among patients with COVID-19 according to the presence and number of comorbidities.

**Results:** Of the 1,590 cases, the mean age was 48.9 years. 686 patients (42.7%) were females. 647 (40.7%) patients were managed inside Hubei province, and 1,334 (83.9%) patients had a contact history of Wuhan city. Severe cases accounted for 16.0% of the study population. 131 (8.2%) patients reached to the composite endpoints. 399 (25.1%) reported having at least one comorbidity. 269 (16.9%), 59 (3.7%), 30 (1.9%), 130 (8.2%), 28 (1.8%), 24 (1.5%), 21 (1.3%), 18 (1.1%) and 3 (0.2%) patients reported having hypertension, cardiovascular diseases, cerebrovascular diseases, diabetes, hepatitis B infections, chronic obstructive pulmonary disease, chronic kidney diseases, malignancy and immunodeficiency, respectively. 130 (8.2%) patients reported having two or more comorbidities. Patients with two or more comorbidities had significantly escalated risks of reaching to the composite endpoint compared with those who had a single comorbidity, and even more so as compared with those without (all *P*<0.05). After adjusting for age and smoking status, patients with COPD (HR 2.681, 95%CI 1.424-5.048), diabetes (HR 1.59, 95%CI 1.03-2.45), hypertension (HR 1.58, 95%CI 1.07-2.32) and malignancy (HR 3.50, 95%CI 1.60-7.64) were more likely to reach to the composite endpoints than those without. As compared with patients without comorbidity, the HR (95%CI) was 1.79 (95%CI 1.16-2.77) among patients with at least one comorbidity and 2.59 (95%CI 1.61-4.17) among patients with two or more comorbidities.

**Conclusion:** Comorbidities are present in around one fourth of patients with COVID-19 in China, and predispose to poorer clinical outcomes.

**Highlights:** *What is already known on this topic:*

- Since November 2019, the rapid outbreak of coronavirus disease 2019 (COVID-19) has recently become a public health emergency of international concern. There have been 79,331 laboratory-confirmed cases and 2,595 deaths globally as of February 25^th^, 2020
- Previous studies have demonstrated the association between comorbidities and other severe acute respiratory diseases including SARS and MERS.
- No study with a nationwide representative cohort has demonstrated the spectrum of comorbidities and the impact of comorbidities on the clinical outcomes in patients with COVID-19.

*What this study adds:*

- In this nationwide study with 1,590 patients with COVID-19, comorbidities were identified in 399 patients. Comorbidities of COVID-19 mainly included hypertension, cardiovascular diseases, cerebrovascular diseases, diabetes, hepatitis B infections, chronic obstructive pulmonary disease, chronic kidney diseases, malignancy and immunodeficiency.
- The presence of as well as the number of comorbidities predicted the poor clinical outcomes (admission to intensive care unit, invasive ventilation, or death) of COVID-19.
- Comorbidities should be taken into account when estimating the clinical outcomes of patients with COVID-19 on hospital admission.

## Introduction

Since November 2019, the rapid outbreak of coronavirus disease 2019 (COVID-19), which arose from severe acute respiratory syndrome coronavirus 2 (SARS-CoV-2) infection, has recently become a public health emergency of international concern [1]. COVID-19 has contributed to an enormous adverse impact globally. Hitherto, there have been 79,331 laboratory-confirmed cases and 2,595 deaths globally as of February 25^th^, 2020 [2].

The clinical manifestations of COVID-19 are, according to the latest reports [3-8], largely heterogeneous. On admission, 20-51% of patients reported as having at least one comorbidity, with diabetes (10-20%), hypertension (10-15%) and cardiovascular and cerebrovascular diseases (7-40%) being most common [3,4,6]. Previous studies have demonstrated that the presence of any comorbidity has been associated with a 3.4-fold increased risk of developing acute respiratory distress syndrome in patients with H7N9 infection [9]. Similar with influenza [10-14], Severe Acute Respiratory Syndrome coronavirus (SARS-CoV) [15] and Middle East Respiratory Syndrome coronavirus (MERS-CoV) [16-24], COVID-19 more readily predisposed to respiratory failure and death in susceptible patients [4]. Nonetheless, previous studies have been certain limitations in study design including the relatively small sample sizes and single center observations. Studies that address these limitations is needed to explore for the factors underlying the adverse impact of COVID-19.

Our objective was to compare the clinical characteristics and outcomes of patients with COVID-19 by stratification according to the presence and category of comorbidity, thus unraveling the subpopulations with poorer prognosis.

## Methods

### Data sources and data extraction

This was a retrospective cohort study that collected data from patients with COVID-19 throughout China, under the coordination of the National Health Commission which mandated the reporting of clinical information from individual designated hospitals which admitted patients with COVID-19. After careful medical chart review, we compiled the clinical data of laboratory-confirmed hospitalized cases from 575 hospitals between November 21^st^, 2019 and January 31^st^, 2020. The diagnosis of COVID-19 was made based on the *World Health Organization* interim guidance [25]. Confirmed cases denoted the patients whose high-throughput sequencing or real-time reverse-transcription polymerase-chain-reaction (RT-PCR) assay findings for nasal and pharyngeal swab specimens were positive [3]. See *Online Supplement* for details.

The clinical data (including recent exposure history, clinical symptoms and signs, comorbidities, and laboratory findings upon admission) were reviewed and extracted by experienced respiratory clinicians, who subsequently entered the data into a computerized database for further cross-checking. Manifestations on chest X-ray or computed tomography (CT) was summarized by integrating the documentation or description in medical charts and, if available, a further review by our medical staff. Major disagreement of the radiologic manifestations between the two reviewers was resolved by consultation with another independent reviewer. Because disease severity reportedly predicted poorer clinical outcomes of avian influenza [9], patients were classified as having severe or non-severe COVID-19 based on the *American Thoracic Society* guidelines for community-acquired pneumonia because of its global acceptance [26].

Comorbidities were determined based on patient’s self-report on admission. Comorbidities were initially treated as a categorical variable (Yes vs. No), and subsequently classified based on the number (Single vs. Multiple). Furthermore, comorbidities were sorted according to the organ systems (i.e. respiratory, cardiovascular, endocrine). Comorbidities that were classified into the same organ system (i.e. coronary heart disease, hypertension) would be merged into a single category.

The primary endpoint of our study was a composite measure which consisted of the admission to intensive care unit (ICU), or invasive ventilation, or death. This composite measure was adopted because all individual components were serious outcomes of H7N9 infections [9]. Secondary endpoints mainly included the mortality rate, and the time from symptom onset to reaching to the composite endpoints.

### Statistical analysis

Statistical analyses were conducted with SPSS software version 23.0 (Chicago, IL, USA). No formal sample size estimation was made because there has not been any published nationwide data on COVID-19. Nonetheless, our sample size was deemed sufficient to power the statistical analysis given its representativeness of the national patient population. Continuous variables were presented as means and standard deviations or medians and interquartile ranges (IQR) as appropriate, and the categorical variables were presented as counts and percentages. Independent t-test, Kruskal-Wallis test and chi-square test were applied for the comparisons between the two groups as appropriate. Cox proportional hazard regression models were applied to determine the potential risk factors associated with the composite endpoints, with the hazards ratio (HR) and 95% confidence interval (95%CI) being reported.

### Patient and public involvement

No patients were directly involved in our study design, setting the research questions, the interpretation of data, or asked to advise on writing up of the report.

## Results

### Demographic and clinical characteristics

The National Health Commission has issued 11,791 patients with laboratory-confirmed COVID-19 in China as of January 31^st^, 2020. At this time point for data cut-off, our database has included 1,590 cases from 575 hospitals in 31 province/autonomous regions/provincial municipalities (see *Online Supplement* for details). Of these 1,590 cases, the mean age was 48.9 years. 686 patients (42.7%) were females. 647 (40.7%) patients were managed inside Hubei province, and 1,334 (83.9%) patients had a contact history of Wuhan city. The most common symptom was fever on or after hospitalization (88.0%), followed by dry cough (70.2%). Fatigue (42.8%) and productive cough (36.0%) were less common. At least one abnormal chest CT manifestation (including ground-glass opacities, pulmonary infiltrates and interstitial disorders) was identified in more than 70% of patients. Severe cases accounted for 16.0% of the study population. 131 (8.2%) patients reached to the composite endpoints during the study (**Table 1)**.

**Table 1:** Demographics and clinical characteristics of patients with or without any comorbidities.

| Variables | Any comorbidity |  |  |  |
| --- | --- | --- | --- | --- |
|  | Total<br>(n=1590) | No<br>(n=1191) | Yes<br>(n=399) | P<br>Value |
| Age (years) | 48.9±16.3 | 44.8±15.2 | 60.8±13.4 | **<0.001** |
| Incubation period (day) | 3.6±4.2 | 3.7±4.3 | 3.5±3.9 | 0.329 |
| Temperature on admission (°C) | 37.4±0.9 | 37.4±0.9 | 37.3±0.9 | **0.034** |
| Respiratory rate on admission (breath/min) | 21.2±12.0 | 21.2±13.7 | 21.3±4.7 | 0.876 |
| Heart rate on admission (beat/min) | 88.7±14.6 | 88.5±14.7 | 89.2±14.4 | 0.402 |
| Systolic pressure on admission (mmHg) | 126.1±16.4 | 123.5±15.2 | 133.2±17.5 | **<0.001** |
| Diastolic pressure on admission (mmHg) | 79.5±25.6 | 79±28.9 | 80.9±12.6 | 0.22 |
| Highest temperature (°C) | 38.3±1.6 | 38.3±1.1 | 38.2±2.6 | 0.634 |
| **Sex** |  |  |  | 0.241 |
| Male | 904/1578 (57.3) | 667/1182 (56.4) | 237/396 (59.8) |  |
| Female | 674/1578 (42.7) | 515/1182 (43.6) | 159/396 (40.2) |  |
| **Smoking status** |  |  |  | **<0.001** |
| Never/unknown | 1479/1590 (93) | 1127/1191 (94.6) | 352/399 (88.2) |  |
| Former/current | 111/1590 (7) | 64/1191 (5.4) | 47/399 (11.8) |  |
| **Symptoms** |  |  |  |  |
| Fever | 1351/1536 (88) | 1002/1148 (87.3) | 349/388 (89.9) | 0.176 |
| Conjunctival congestion | 10/1345 (0.7) | 7/1014 (0.7) | 3/331 (0.9) | 0.715 |
| Nasal congestion | 73/1299 (5.6) | 59/979 (6) | 14/320 (4.4) | 0.328 |
| Headache | 205/1328 (15.4) | 151/1002 (15.1) | 54/326 (16.6) | 0.537 |
| Dry cough | 1052/1498 (70.2) | 775/1116 (69.4) | 277/382 (72.5) | 0.271 |
| Pharyngodynia | 194/1317 (14.7) | 148/999 (14.8) | 46/318 (14.5) | 0.928 |
| Productive cough | 513/1424 (36) | 363/1064 (34.1) | 150/360 (41.7) | **0.011** |
| Fatigue | 584/1365 (42.8) | 435/1031 (42.2) | 149/334 (44.6) | 0.446 |
| Hemoptysis | 16/1315 (1.2) | 9/991 (0.9) | 7/324 (2.2) | 0.084 |
| Shortness of breath | 331/1394 (23.7) | 185/1041 (17.8) | 146/353 (41.4) | **<0.001** |
| Nausea/vomiting | 80/1371 (5.8) | 44/1025 (4.3) | 36/346 (10.4) | **<0.001** |
| Diarrhea | 57/1359 (4.2) | 39/1023 (3.8) | 18/336 (5.4) | 0.213 |
| Myalgia/arthralgia | 234/1338 (17.5) | 174/1007 (17.3) | 60/331 (18.1) | 0.739 |
| Chill | 163/1333 (12.2) | 129/1006 (12.8) | 34/327 (10.4) | 0.285 |
| **Signs** |  |  |  |  |
| Throat congestion | 21/1286 (1.6) | 16/973 (1.6) | 5/313 (1.6) | 1 |
| Tonsil swelling | 31/1376 (2.3) | 22/1024 (2.1) | 9/352 (2.6) | 0.678 |
| Enlargement of lymph nodes | 2/1375 (0.1) | 1/1027 (0.1) | 1/348 (0.3) | 0.442 |
| Rash | 3/1378 (0.2) | 2/1032 (0.2) | 1/346 (0.3) | 1 |
| Unconsciousness | 20/1421 (1.4) | 11/1063 (1) | 9/358 (2.5) | 0.064 |
| **Abnormal chest image** |  |  |  |  |
| Radiograph | 243/1590 (15.3) | 236/1566 (15.1) | 44036 (29.2) | 0.079 |
| Computed tomography | 1130/1590 (71.1) | 1113/1566 (71.1) | 17/24 (70.8) | 1 |
| **Hubei** |  |  |  | **<0.001** |
| Yes | 647/1590 (40.7) | 434/1191 (36.4) | 213/399 (53.4) |  |
| No | 943/1590 (59.3) | 757/1191 (63.6) | 186/399 (46.6) |  |
| **Wuhan-contacted** |  |  |  | **0.012** |
| Yes | 1334/1590 (83.9) | 983/1191 (82.5) | 351/399 (88) |  |
| No | 256/1590 (16.1) | 208/1191 (17.5) | 48/399 (12) |  |
| **Severity** | 254/1590 (16) | 123/1191 (10.3) | 131/399 (32.8) | **<0.001** |
| **Composite endpoint** | 131/1590 (8.2) | 54/1191 (4.5) | 77/399 (19.3) | **<0.001** |
| **Death** | 50/1590 (3.1) | 15/1191 (1.3) | 35/399 (8.8) | **<0.001** |

Data are mean ± standard deviation, n/N (%), where N is the total number of patients with available data. p values are calculated by $\chi^2$ test, Fisher's exact test, or Mann-Whitney U test. COPD=chronic obstructive pulmonary disease.
Data in bold indicated the statistical comparisons with significance.

### Presence of comorbidities and the clinical characteristics and outcomes of COVID-19

Of the 1,590 cases, 399 (25.1%) reported having at least one comorbidity. The most common comorbidities encompassed hypertension (269 [16.9%]), diabetes (130 [8.2%]), and cardiovascular diseases (59 [3.7%]). Chronic obstructive pulmonary disease (COPD) was identified in 24 cases. At least one comorbidity was seen more commonly in severe cases than in non-severe cases (32.8% vs. 10.3%). Patients with at least one comorbidity were older (mean: 60.8 vs. 44.8 years), were more likely to have shortness of breath (41.4% vs. 17.8%), nausea or vomiting (10.4% vs. 4.3%), and tended to have abnormal chest X-ray manifestations (29.2% vs. 15.1%) (**Table 1)**.

### Clinical characteristics and outcomes of COVID-19 stratified by the number of comorbidities

We have further identified 130 (8.2%) patients who reported having two or more comorbidities. Two or more comorbidities were more commonly seen in severe cases than in non-severe cases (40.0% vs. 29.4%, *P*<0.001). Patients with two or more comorbidities were older (mean: 66.2 vs. 58.2 years), were more likely to have shortness of breath (55.4% vs. 34.1%), nausea or vomiting (11.8% vs. 9.7%), unconsciousness (5.1% vs. 1.3%) and less abnormal chest X-ray (20.8% vs. 23.4%) compared with patients who had single comorbidity (**Table 2)**.

**Table 2:** Demographics and clinical characteristics of patients with 1 or ≥2 comorbidities.

| **Variables** | **1 comorbidity (n=269)** | **$\geq 2$ comorbidities (n=130)** | **P Value** |
| --- | --- | --- | --- |
| Age (years) | 58.2±13.1 | 66.2±12.2 | **<0.001** |
| Incubation period (days) | 3.2±3.1 | 4.0±5.2 | 0.124 |
| Temperature on admission ($^{\circ}$C) | 37.4±0.9 | 37.1±0.9 | **<0.001** |
| Respiratory rate on admission (breath/min) | 21.4±4.6 | 21.2±5 | 0.977 |
| Heart rate (bit/minute) | 90.2±14.6 | 87.2±13.7 | 0.134 |
| Systolic pressure on admission (mmHg) | 132.2±16.5 | 135.3±19.4 | **<0.001** |
| Diastolic pressure on admission (mmHg) | 81.7±12.5 | 79.5±12.9 | 0.350 |
| Highest temperature ($^{\circ}$C) | 38.2±3.0 | 38.4±0.8 | 0.424 |
| **Sex** |  |  | 0.430 |
| Male | 158/268 (59.0) | 79/128 (61.7) |  |
| Female | 110/268 (41.0) | 49/128 (38.3) |  |
| **Smoking status** |  |  | **<0.001** |
| Never/unknown | 234/269 (87.0) | 118/130 (90.8) |  |
| Former/current | 35/269 (13.0) | 12/130 (9.2) |  |
| **Symptoms** |  |  |  |
| Fever | 241/263 (91.6) | 108/125 (86.4) | 0.126 |
| Conjunctival congestion | 3/222 (1.4) | 0/109 (0) | 0.374 |
| Nasal congestion | 5/213 (2.3) | 9/107 (8.4) | **0.046** |
| Headache | 34/220 (15.5) | 20/106 (18.9) | 0.589 |
| Dry cough | 195/258 (75.6) | 82/124 (66.1) | 0.088 |
| Pharyngodynia | 33/218 (15.1) | 13/100 (13.0) | 0.872 |
| Productive cough | 101/241 (41.9) | 49/119 (41.2) | **0.036** |
| Fatigue | 97/227 (42.7) | 52/107 (48.6) | 0.444 |
| Hemoptysis | 4/219 (1.8) | 3/105 (2.9) | 0.149 |
| Shortness of breath | 79/232 (34.1) | 67/121 (55.4) | <0.001 |
| Nausea/vomiting | 23/236 (9.7) | 13/110 (11.8) | **<0.001** |
| Diarrhea | 11/229 (4.8) | 7/107 (6.5) | 0.359 |
| Myalgia/arthralgia | 45/227 (19.8) | 15/104 (14.4) | 0.457 |
| Chill | 25/222 (11.3) | 9/105 (8.6) | 0.400 |
| **Signs** |  |  |  |
| Throat congestion | 4/216 (1.9) | 1/97 (1) | 0.868 |
| Tonsil swelling | 5/234 (2.1) | 4/118 (3.4) | 0.685 |
| Enlargement of lymph nodes | 1/232 (0.4) | 0/116 (0) | 0.441 |
| Rash | 0/231 (0) | 1/115 (0.9) | 0.249 |
| Unconsciousness | 3/240 (1.3) | 6/118 (5.1) | **0.002** |
| **Abnormal chest image** |  |  |  |
| Radiograph | 63/269 (23.4) | 27/130 (20.8) | **<0.001** |
| Computed tomography | 200/269 (74.3) | 96/130 (73.8) | 0.283 |
| **Hubei** |  |  | **<0.001** |
| Yes | 120/269 (44.6) | 93/130 (71.5) |  |
| No | 149/269 (55.4) | 37/130 (28.5) |  |
| **Wuhan-contacted** |  |  | **0.003** |
| Yes | 229/269 (85.1) | 122/130 (93.8) |  |
| No | 40/269 (14.9) | 8/130 (6.2) |  |
| **Severity** | 79/269 (29.4) | 52/130 (40.0) | **<0.001** |
| **Composite endpoint** | 40/269 (14.9) | 37/130 (28.5) | **<0.001** |
| **Deaths** | 15/269 (5.6) | 20/130 (15.4) | **<0.001** |

Data are mean ± standard deviation, n/N (%), where N is the total number of patients with available data. p values are calculated by $\chi^2$ test, Fisher's exact test, or Mann-Whitney U test. COPD=chronic obstructive pulmonary disease.
Data in bold indicated the statistical comparisons with significance.

### Clinical characteristics and outcomes of COVID-19 stratified by organ systems of comorbidities

A total of 269 (16.9%), 59 (3.7%), 30 (1.9%), 130 (8.2%), 28 (1.8%), 24 (1.5%), 21 (1.3%), 18 (1.1%) and 3 (0.2%) patients reported having hypertension, cardiovascular diseases, cerebrovascular diseases, diabetes, hepatitis B infections, COPD, chronic kidney diseases, malignancy and immunodeficiency, respectively. Severe cases were more likely to have hypertension (32.7% vs. 12.6%), cardiovascular diseases (33.9% vs. 15.3%), cerebrovascular diseases (50.0% vs. 15.3%), diabetes (34.6% vs. 14.3%), hepatitis B infections (32.1% vs. 15.7%), COPD (62.5% vs. 15.3%), chronic kidney diseases (38.1% vs. 15.7%) and malignancy (50.0% vs. 15.6%) compared with non-severe cases. Furthermore, comorbidities were more common patients treated in Hubei province as compared with those managed outside Hubei province (all *P*<0.05) as well as patients with an exposure history of Wuhan as compared with those without (all *P*<0.05) (**Table 3)**.

**Table 3:**
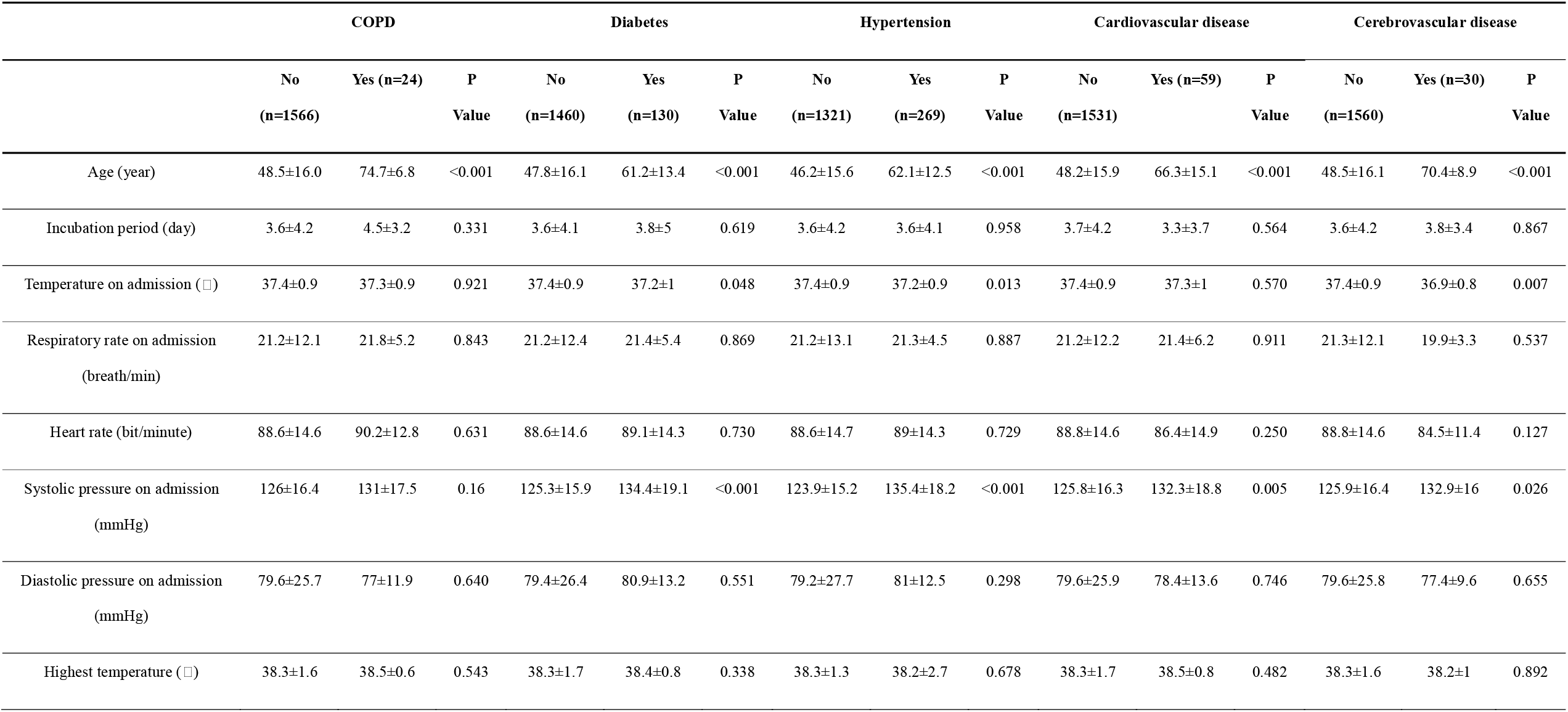
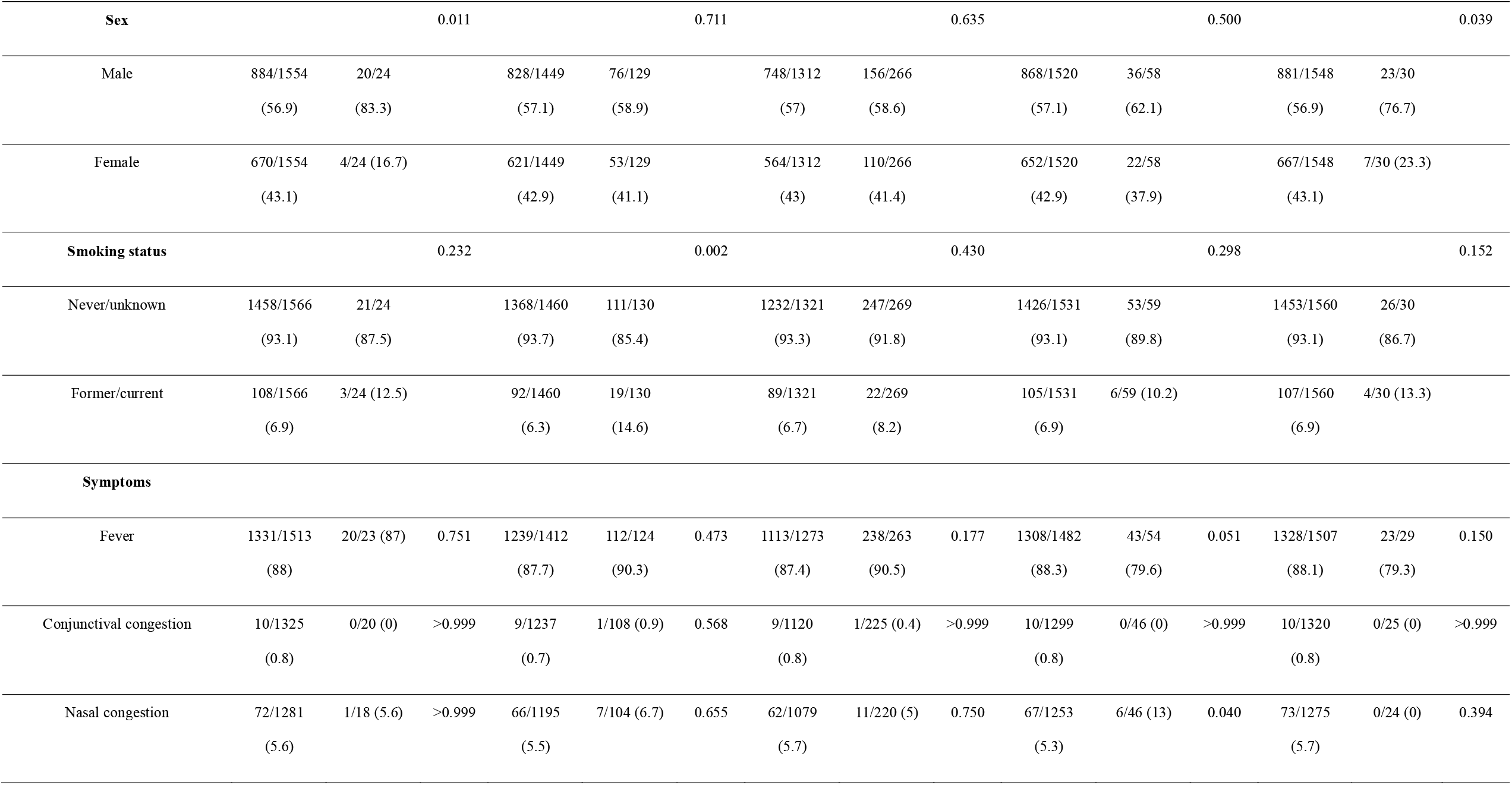
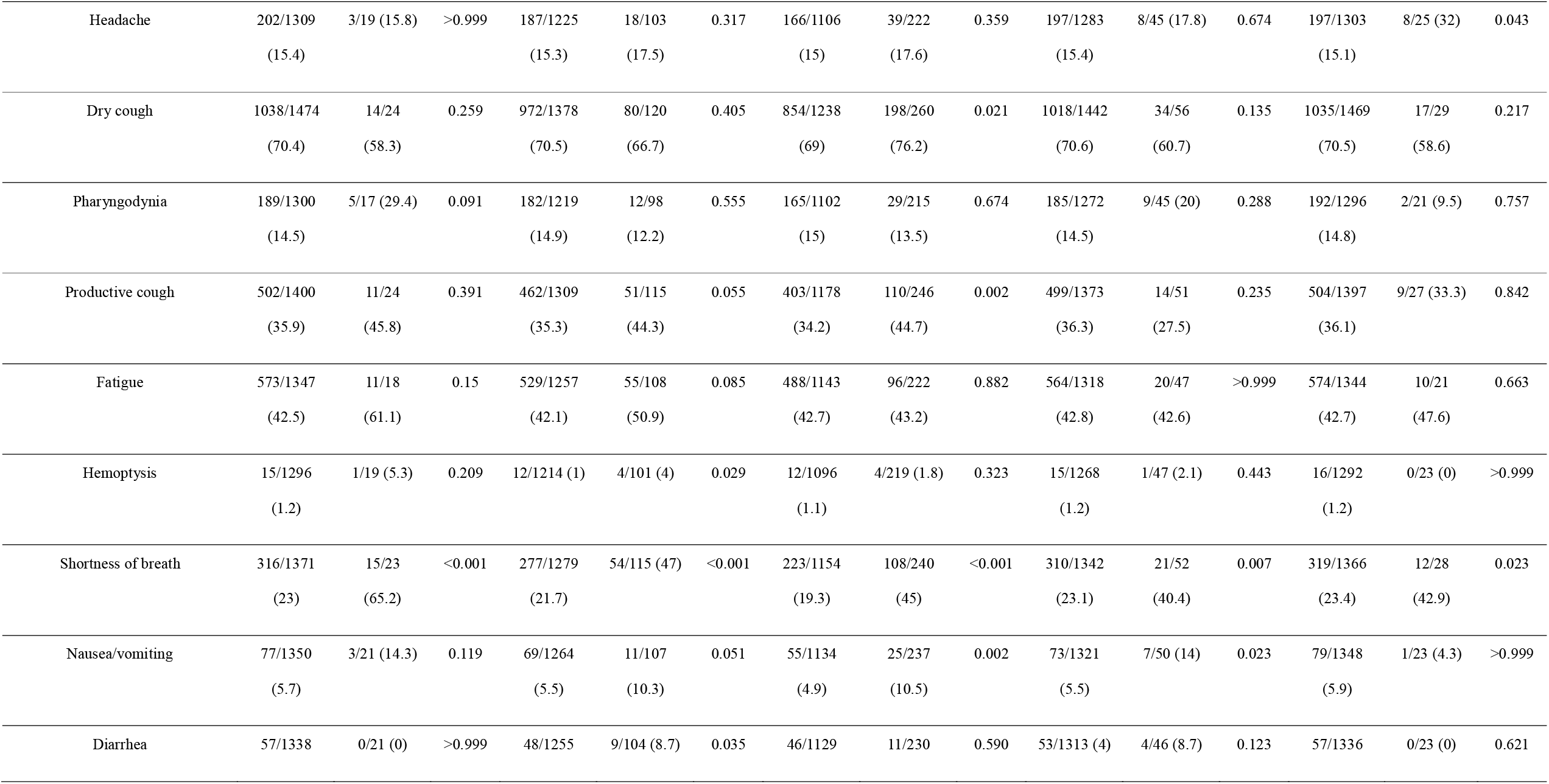
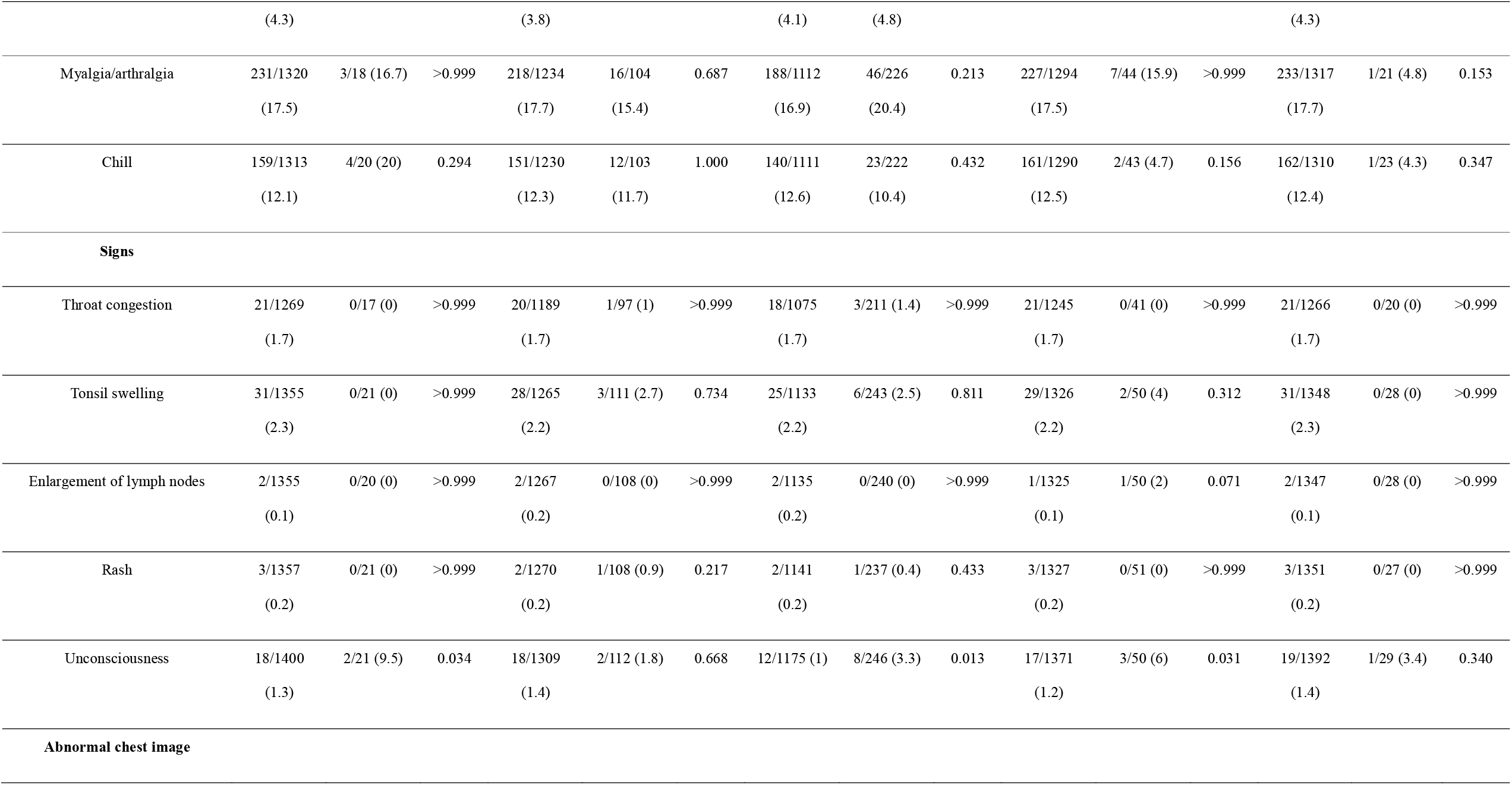
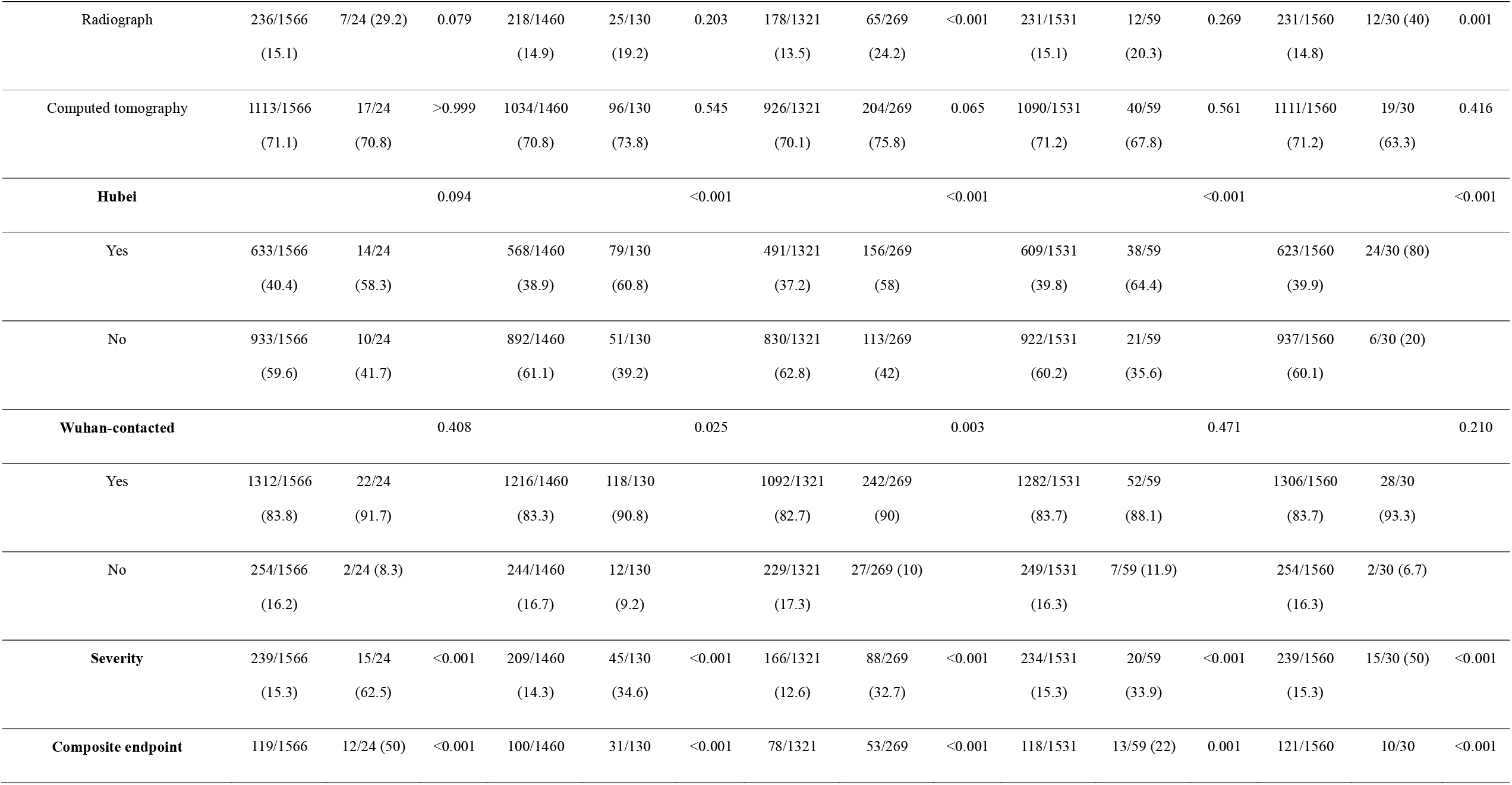
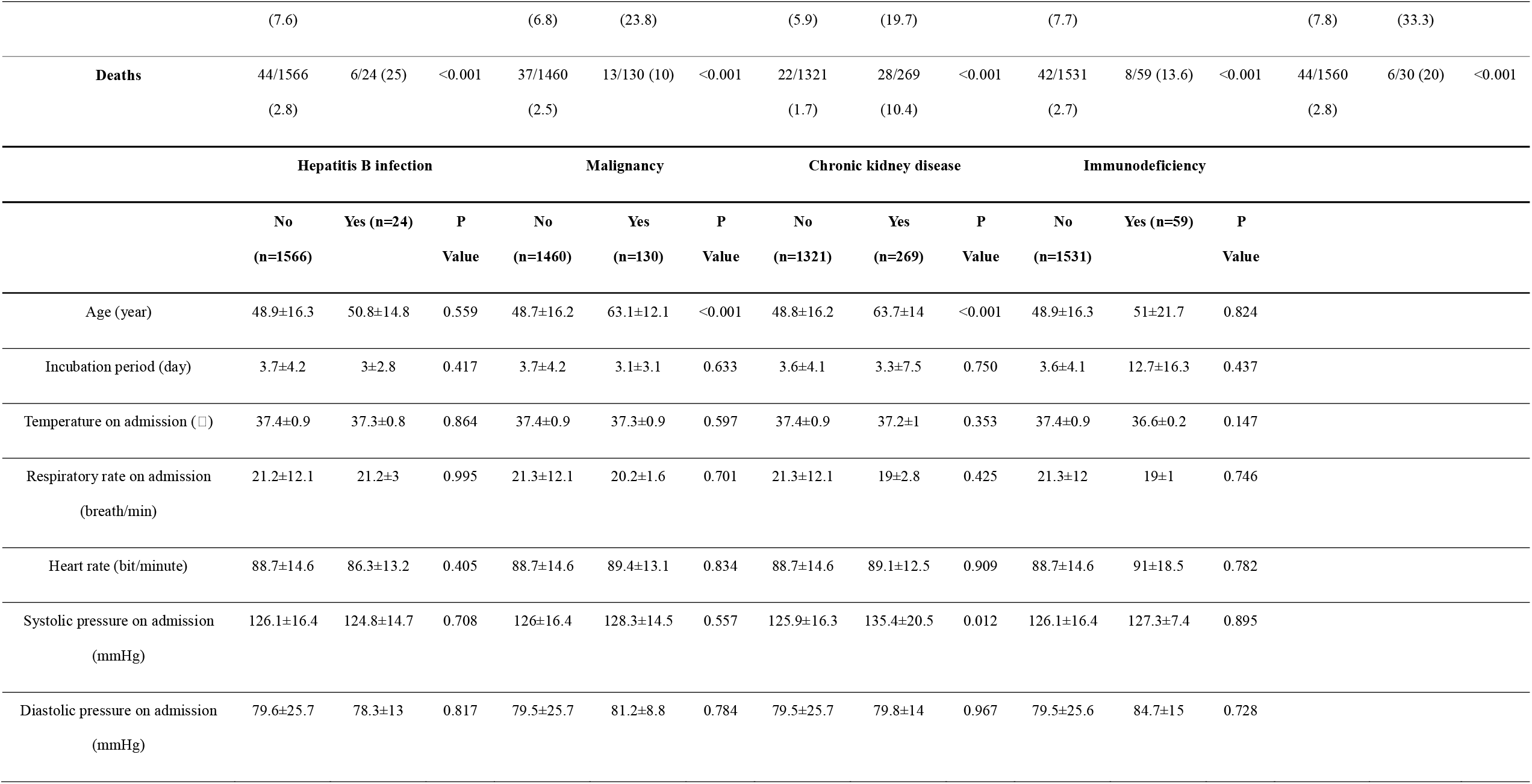
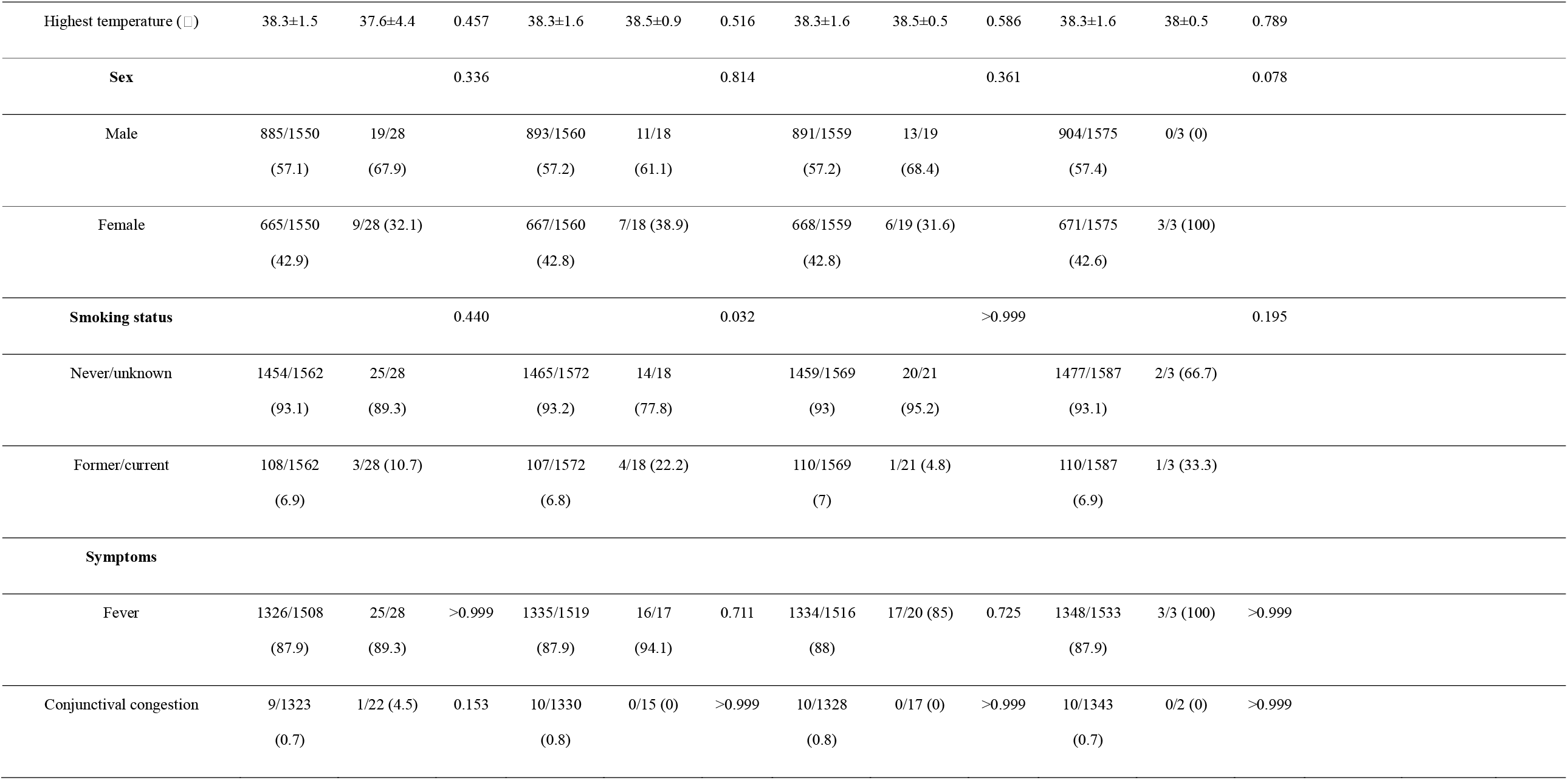
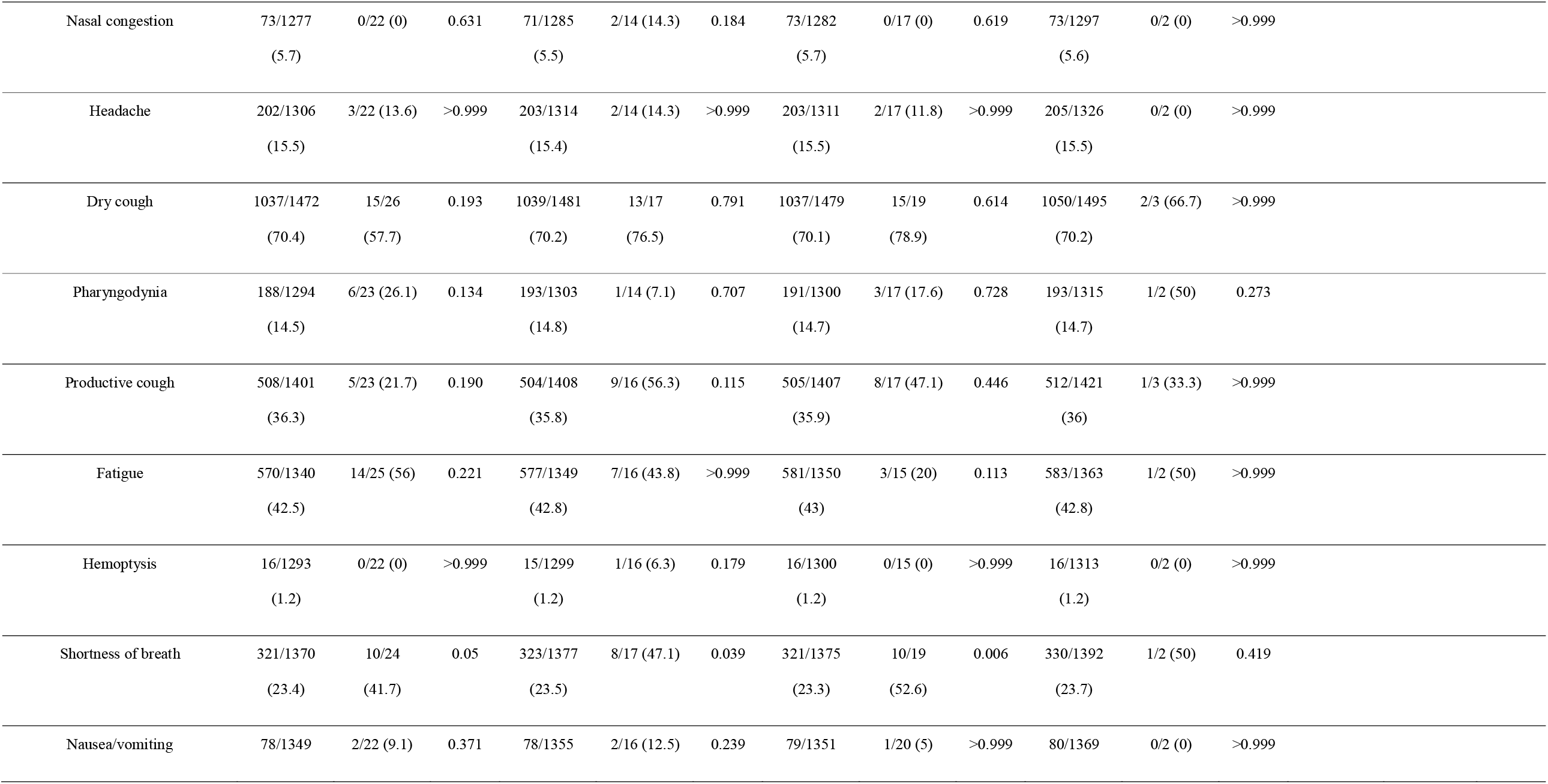
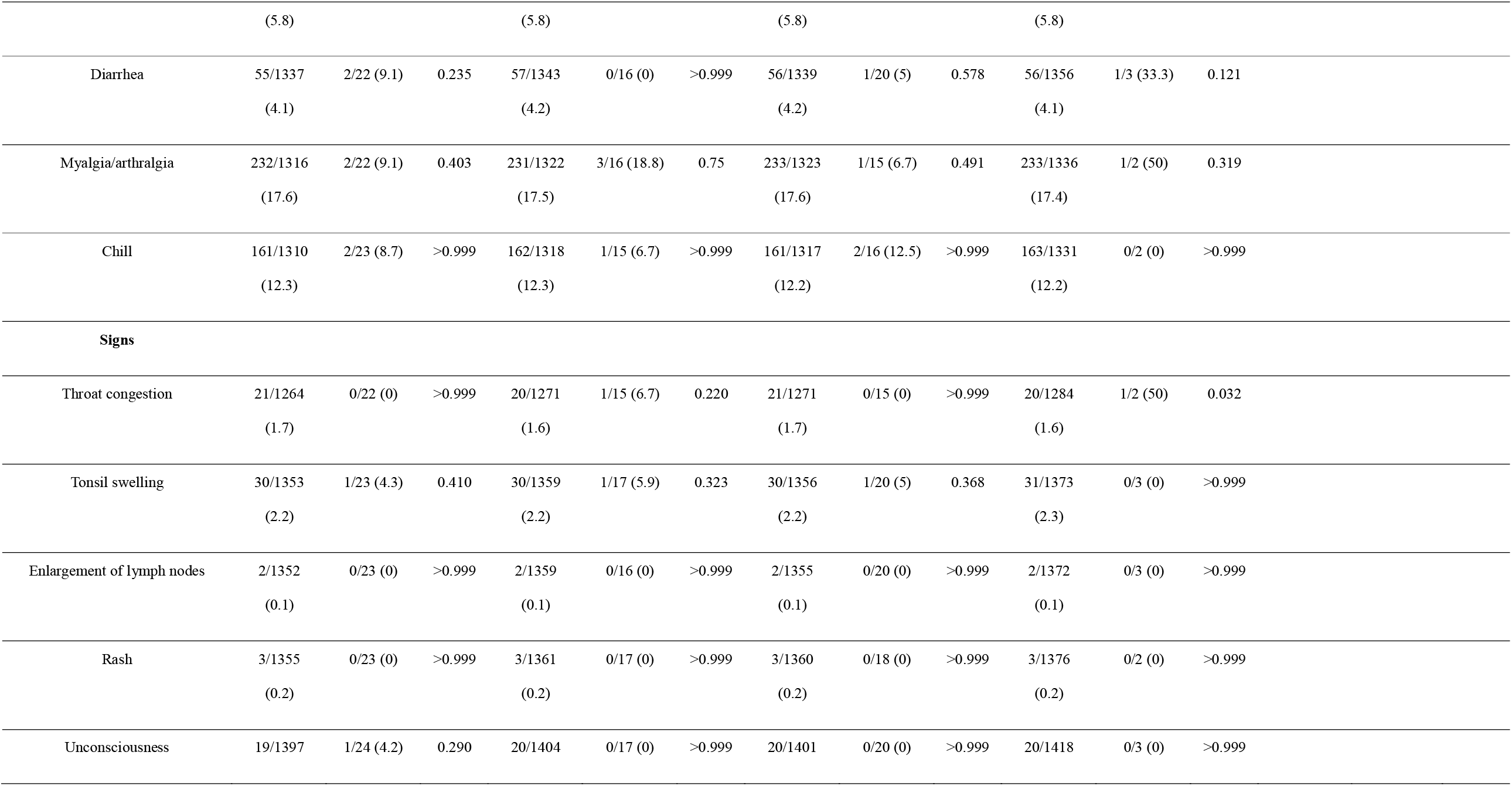
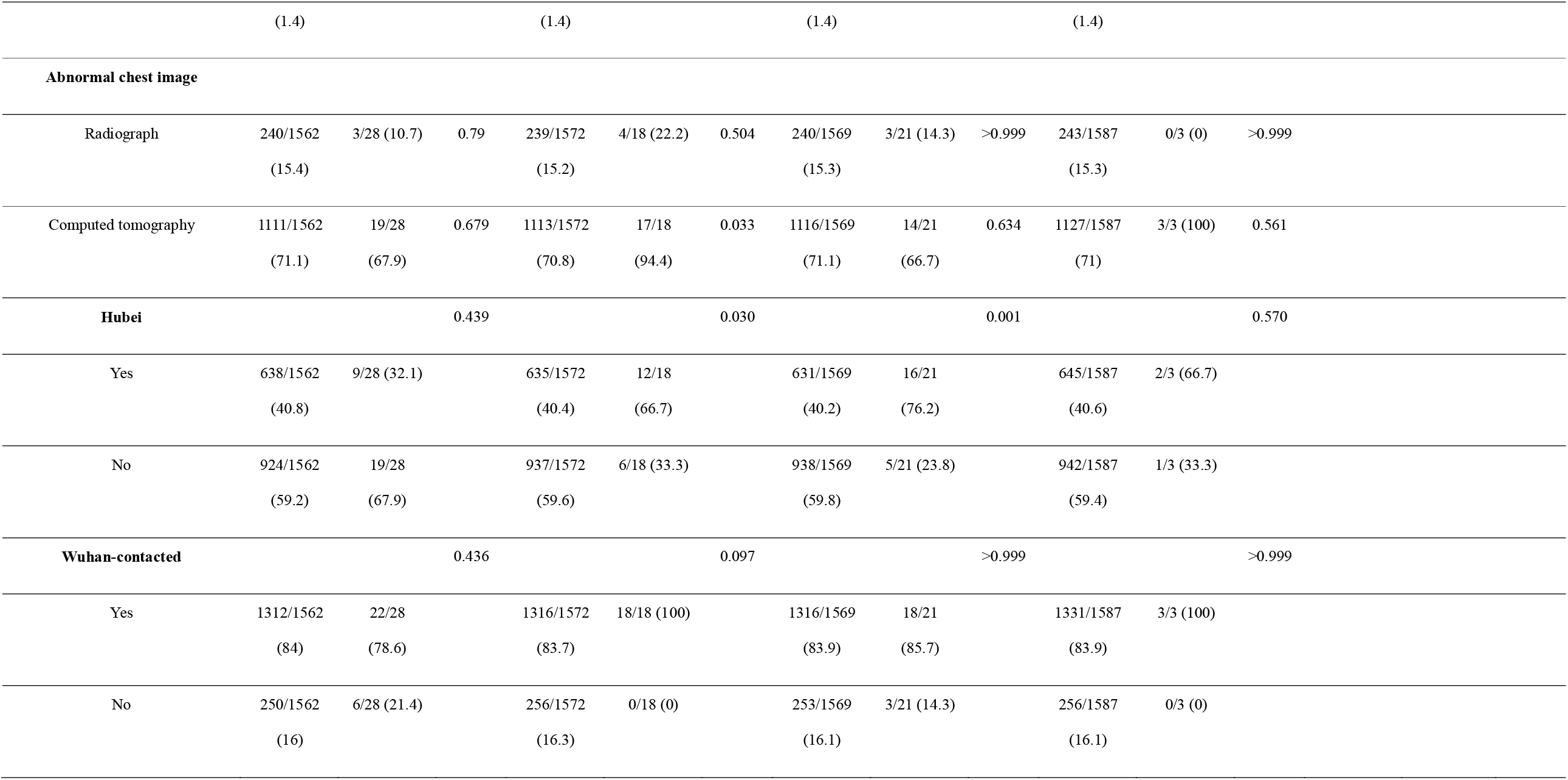
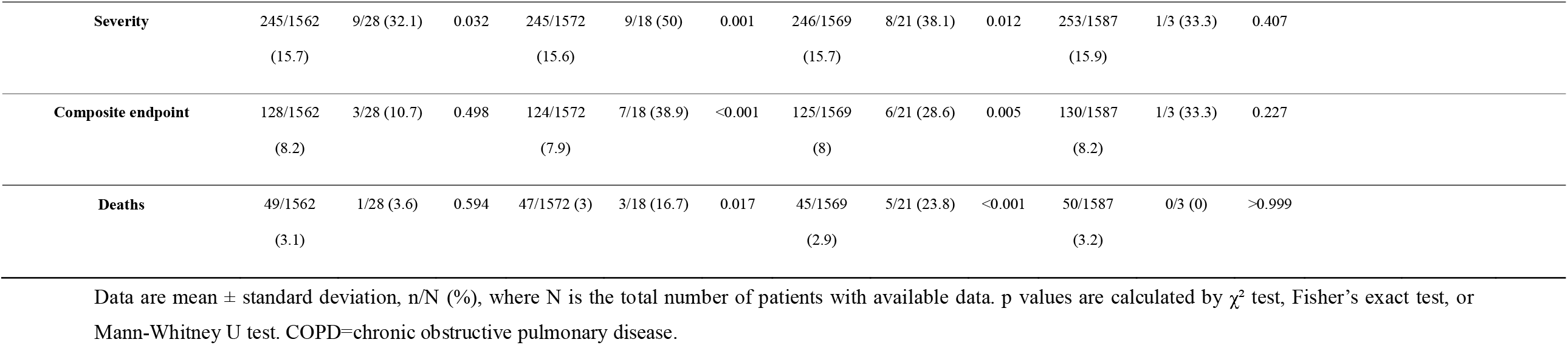
Demographics and clinical characteristics of patients stratified by different comorbidities.

### Prognostic analyses

The composite endpoint was documented in 77 (19.3%) of patients who had at least one comorbidity as opposed to 54 (4.5%) patients without comorbidities (*P*<0.001). This figure was 37 cases (28.5%) in patients who had two or more comorbidities. Significantly more patients with hypertension (19.7% vs. 5.9%), cardiovascular diseases (22.0% vs. 7.7%), cerebrovascular diseases (33.3% vs. 7.8%), diabetes (23.8% vs. 6.8%), COPD (50.0% vs. 7.6%), chronic kidney diseases (28.6% vs. 8.0%) and malignancy (38.9% vs. 7.9%) reached to the composite endpoints compared with those without (**Table 3**).

Patients with two or more comorbidities had significantly escalated risks of reaching to the composite endpoint compared with those who had a single comorbidity, and even more so as compared with those without (all *P*<0.05, **Figure 1**). After adjusting for age and smoking status, patients with COPD (HR 2.68, 95%CI 1.42-5.05), diabetes (HR 1.59, 95%CI 1.03-2.45), hypertension (HR 1.58, 95%CI 1.07-2.32) and malignancy (HR 3.50, 95%CI 1.60-7.64) were more likely to reach to the composite endpoints than those without (**Figure 2**). As compared with patients without comorbidity, the HR (95%CI) was 1.79 (95%CI 1.16-2.77) among patients with at least one comorbidity and 2.59 (95%CI 1.61-4.17) among patients with two or more comorbidities (**Figure 2**).

**Figure 1.**
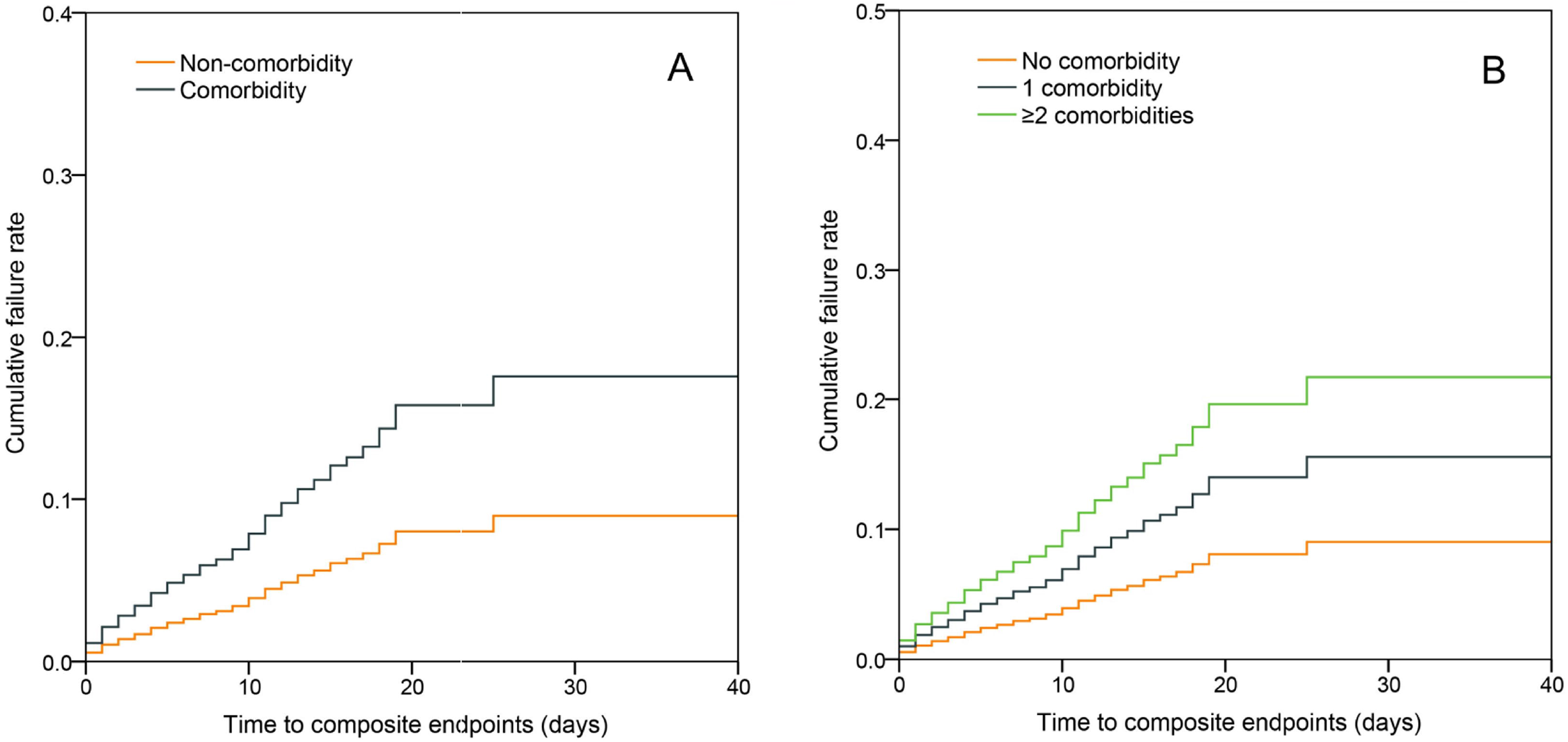
Comparison of the time-dependent risk of reaching to the composite endpoints. Figure 1-A, The time-dependent risk of reaching to the composite endpoints between patients with (orange curve) or without any comorbidity (dark blue curve); Figure 1-B, The time-dependent risk of reaching to the composite endpoints between patients without any comorbidity (orange curve), patients with a single comorbidity (dark blue curve), and patients with two or more comorbidities (green curve).

**Figure 2.**
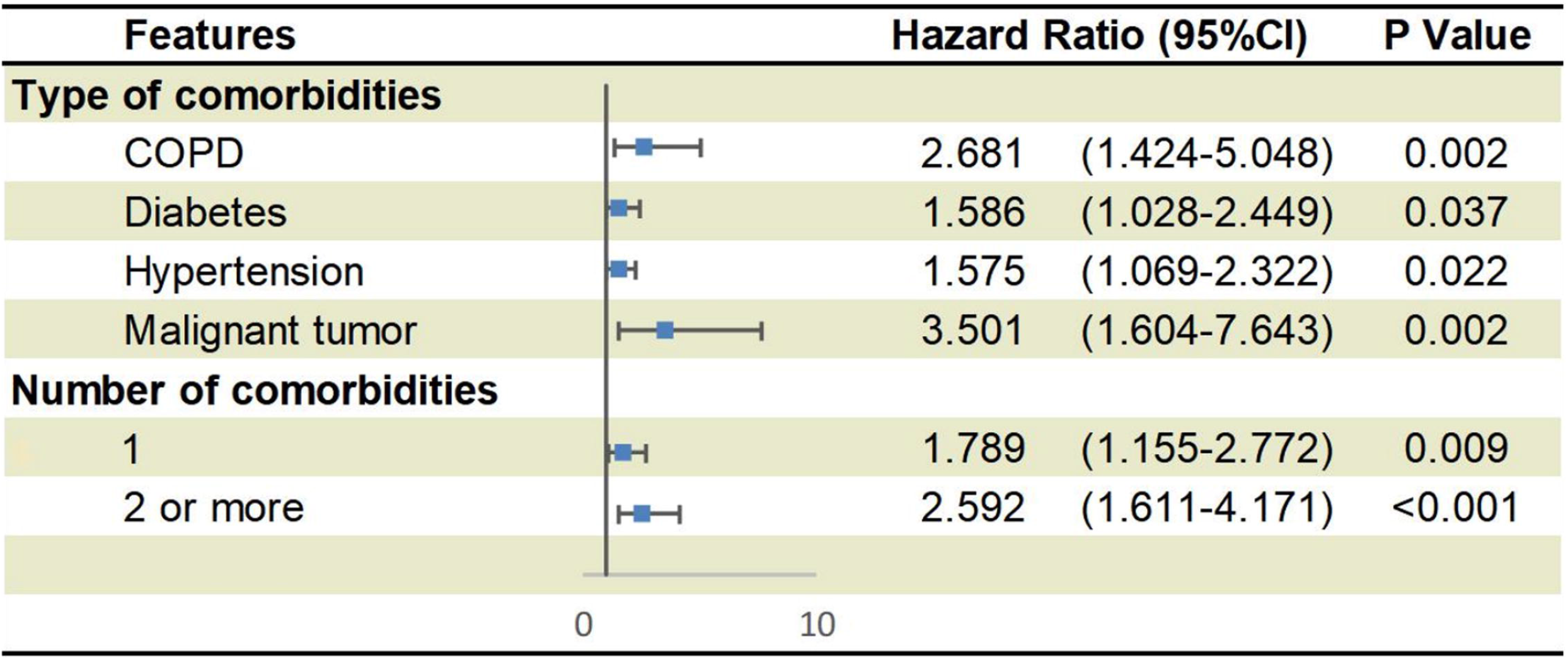
Predictors of the composite endpoints in the proportional hazards model. Shown in the figure are the hazards ratio (HR) and the 95% confidence interval (95%CI) for the risk factors associated with the composite endpoints (admission to intensive care unit, invasive ventilation, or death). The comorbidities were classified according to the organ systems as well as the number. The scale bar indicates the HR. The model has been adjusted with age and smoking status

## Discussion

Our study is the first nationwide investigation that systematically evaluates the impact of comorbidities on the clinical characteristics and prognosis in patients with COVID-19 in China. Circulatory and endocrine comorbidities were common among patients with COVID-19. Patients with at least one comorbidity, or more even so, were associated with poor clinical outcomes. These findings have provided further objective evidence, with a large sample size and extensive coverage of the geographic regions across China, to take into account baseline comorbid diseases in the comprehensive risk assessment of prognosis among patients with COVID-19 on hospital admission.

Overall, our findings have echoed the recently published studies in terms of the commonness of comorbidities in patients with COVID-19 [3-7]. Despite considerable variations in the proportion in individual studies due to the limited sample size and the region where patients were managed, circulatory diseases (including hypertension and coronary heart diseases) remained the most common category of comorbidity [3-7]. Apart from circulatory diseases, endocrine diseases such as diabetes were also common in patients with COVID-19. Notwithstanding the commonness of circulatory and endocrine comorbidities, patients with COVID-19 rarely reported as having comorbid respiratory diseases (particularly COPD). The reasons underlying this observation have been scant, but could have arisen from the lack of awareness and the lack of spirometric testing in community settings that collectively contributed to the underdiagnosis of respiratory diseases [27]. Consistent with recent reports [3-7], the percentage of patients with comorbid renal disease and malignancy was relatively low. Our findings have therefore added to the existing literature the spectrum of comorbidities in patients with COVID-19 based on the larger sample sizes and representativeness of the whole patient population in China.

A number of existing literature reports have documented the escalated risks of poorer clinical outcomes in patients with avian influenza [10-14], SARS-CoV [15] and MERS-CoV infections [16-24]. The most common comorbidities associated with poorer prognosis included diabetes [21,24], hypertension [24], respiratory diseases [15,24], cardiac diseases [15,24], pregnancy [12], renal diseases [24] and malignancy [15]. Our findings suggested that, similar with other severe acute respiratory outbreaks, comorbidities such as COPD, diabetes, hypertension and malignancy predisposed to adverse clinical outcomes in patients with COVID-19. The strength of association between different comorbidities and the prognosis, however, was less consistent when compared with the literature reports [12,15,21,24]. For instance, the risk between cardiac diseases and poor clinical outcomes of influenza, SARS-CoV or MERS-CoV infections was inconclusive [12,15,21,24]. Except for diabetes, no other comorbidities were identified to be the predictors of poor clinical outcomes in patients with MERS-CoV infections [21]. Few studies, however, have explored the mechanisms underlying these associations. Kulscar et al showed that MERS-CoV infections resulted in prolonged airway inflammation, immune cell dysfunction and an altered expression profile of inflammatory mediators [23]. A network-based analysis indicated that SARS-CoV infections led to immune dysregulation that could help explain the escalated risk of cardiac diseases, bone diseases and malignancy [28]. Therefore, immune dysregulation and prolonged inflammation might be the key drivers of the poor clinical outcomes in patients with COVID-19 but await verification in more mechanistic studies.

There has been a considerable overlap in the comorbidities which has been widely accepted. For instance, diabetes [29] and COPD [30] frequently co-exist with hypertension or coronary heart diseases. Therefore, patients with co-existing comorbidities are more likely to have poorer baseline well-being. Importantly, we have verified the significantly escalated risk of poor prognosis in patients with two or more comorbidities as compared with those who had no or only a single comorbidity. Our findings implied that both the category and number of comorbidities should be taken into account when predicting the prognosis in patients with COVID-19.

Our findings suggested that patients with comorbidities had greater disease severity compared with those without. A greater number of comorbidities correlated with greater disease severity of COVID-19. The public health implication of our study was that proper triage of patients should be implemented in out-patient clinics or on hospital admission by carefully inquiring the medical history because this will help identify patients who would be more likely to develop serious adverse outcomes during the progression of COVID-19. A multidisciplinary team with specialists would be needed to manage the comorbid conditions in a timely fashion. Moreover, patients with COIVD-19 who had comorbidities should be isolated immediately upon confirmation of the diagnosis, which would help provide with this susceptible population better personal medical protection.

The main limitation of our study was the self-report of comorbidities on admission. Underreporting of comorbidities, which could have stemmed from the lack of awareness and/or the lack of diagnostic testing, might contribute to the underestimation of the true strength of association with the clinical prognosis. However, significant underreporting was unlikely because the spectrum of our report was largely consistent with existing literature [3-7] and all patients were subject to a thorough history taking after hospital admission. Moreover, the duration of follow-up was relatively short and some patients remained in the hospital as of the time of writing. More studies that explore the associations in a sufficiently long time frame are warranted. As with other observational studies, our findings did not provide direct inference about the causation or reverse causation of comorbidities and the poor clinical outcomes.

## Conclusions

Comorbidities are present in around one fourth of patients with COVID-19 in China, and predispose to poorer clinical outcomes. A thorough assessment of comorbidities may help establish risk stratification of patients with COVID-19 upon hospital admission.

## Data Availability

Data would be available upon request to Prof Zhong

## Acknowledgment

We thank the hospital staff (see Supplementary Appendix for the full list) for their efforts in collecting the information. We are indebted to the coordination of Drs. Zong-jiu Zhang, Ya-hui Jiao, Bin Du, Xin-qiang Gao and Tao Wei (National Health Commission), Yu-fei Duan and Zhi-ling Zhao (Health Commission of Guangdong Province), Yi-min Li, Zi-jing Liang, Nuo-fu Zhang, Shi-yue Li, Qing-hui Huang, Wen-xi Huang and Ming Li (Guangzhou Institute of Respiratory Health) which greatly facilitate the collection of patient’s data. Special thanks are given to the statistical team members Prof. Zheng Chen, Drs. Dong Han, Li Li, Zheng Chen, Zhi-ying Zhan, Jin-jian Chen, Li-jun Xu, Xiao-han Xu (State Key Laboratory of Organ Failure Research, Department of Biostatistics, Guangdong Provincial Key Laboratory of Tropical Disease Research, School of Public Health, Southern Medical University). We also thank Li-qiang Wang, Wei-peng Cai, Zi-sheng Chen (the sixth affiliated hospital of Guangzhou medical university), Chang-xing Ou, Xiao-min Peng, Si-ni Cui, Yuan Wang, Mou Zeng, Xin Hao, Qi-hua He, Jing-pei Li, Xu-kai Li, Wei Wang, Li-min Ou, Ya-lei Zhang, Jing-wei Liu, Xin-guo Xiong, Wei-juna Shi, San-mei Yu, Run-dong Qin, Si-yang Yao, Bo-meng Zhang, Xiao-hong Xie, Zhan-hong Xie, Wan-di Wang, Xiao-xian Zhang, Hui-yin Xu, Zi-qing Zhou, Ying Jiang, Ni Liu, Jing-jing Yuan, Zheng Zhu, Jie-xia Zhang, Hong-hao Li, Wei-hua Huang, Lu-lin Wang, Jie-ying Li, Li-fen Gao, Jia-bo Gao, Cai-chen Li, Xue-wei Chen, Jia-bo Gao, Ming-shan Xue, Shou-xie Huang, Jia-man Tang, Wei-li Gu, Jin-lin Wang (Guangzhou Institute of Respiratory Health) for their dedication to data entry and verification. We are grateful to Tecent Co. Ltd. for their provision of the number of certified hospitals for admission of patients with COVID-19 throughout China. Finally, we thank all the patients who consented to donate their data for analysis and the medical staffs working in the front line.

## Statements

All authors have completed the ICMJE uniform disclosure form at http://www.icmje.org/coi_disclosure.pdf and declare: no support from any organisation for the submitted work; no financial relationships with any organisations that might have an interest in the submitted work in the previous three years, no other relationships or activities that could appear to have influenced the submitted work.

The Corresponding Author has the right to grant on behalf of all authors and does grant on behalf of all authors, a worldwide licence (http://www.bmj.com/sites/default/files/BMJ%20Author%20Licence%20March%202013.doc) to the Publishers and its licensees in perpetuity, in all forms, formats and media (whether known now or created in the future), to i) publish, reproduce, distribute, display and store the Contribution, ii) translate the Contribution into other languages, create adaptations, reprints, include within collections and create summaries, extracts and/or, abstracts of the Contribution and convert or allow conversion into any format including without limitation audio, iii) create any other derivative work(s) based in whole or part on the on the Contribution, iv) to exploit all subsidiary rights to exploit all subsidiary rights that currently exist or as may exist in the future in the Contribution, v) the inclusion of electronic links from the Contribution to third party material where-ever it may be located; and, vi) licence any third party to do any or all of the above. All research articles will be made available on an open access basis (with authors being asked to pay an open access fee—seehttp://www.bmj.com/about-bmj/resources-authors/forms-policies-and-checklists/copyright-open-access-and-permission-reuse). The terms of such open access shall be governed by a Creative Commons licence—details as to which Creative Commons licence will apply to the research article are set out in our worldwide licence referred to above.

## Author’s contributions

W. J. G., W. H. L., J. X. H., and N. S. Z. participated in study design and study conception; W. H. L., Y. Z., H. R. L., Z. S. C., C. Q. O., L. L., P. Y. C., J. F. L., C. C. L., L. M. O., B. C., W. W. and S. X. performed data analysis; R. C. C., C. L. T., T. W., L. S., Z. Y. N., J. X., Y. H., L. L., H. S., C. L. L., Y. X. P., L. W., Y. L., Y. H. H., P. P., J. M. W., J. Y. L., Z. C., G. L., Z. J. Z., S. Q. Q., J. L., C. J. Y., S. Y. Z., L. L. C., F. Y., S. Y. L., J. P. Z., N. F. Z., and N. S. Z. recruited patients; W. J. G., J. X. H., W. H. L., and N. S. Z. drafted the manuscript; all authors provided critical review of the manuscript and approved the final draft for publication.

## Reference

1. WHO main website. https://www.who.int (accessed February 25th, 2020)

2. World Health Organization. Novel Coronavirus (2019-nCoV) situation reports. https://www.who.int/emergencies/diseases/novel-coronavirus-2019/situation-reports/ (Assessed on February 25th, 2020)

3. Huang C, Wang Y, Li X, et al. Clinical features of patients with 2019 novel coronavirus in Wuhan, China. Lancet. 2020; doi: 10.1016/S0140-6736(20)30183-5

4. Chen N, Zhou M, Dong X, et al. Epidemiological and clinical characteristics of 99 cases of 2019 novel coronavirus pneumonia in Wuhan, China: a descriptive study. Lancet. 2020. doi: 10.1016/S0140-6736(20)30211-7

5. Wang D, Hu B, Hu C, et al. Clinical Characteristics of 138 Hospitalized Patients With 2019 Novel Coronavirus-Infected Pneumonia in Wuhan, China. JAMA. 2020 Feb 7. doi: 10.1001/jama.2020.1585

6. Kui L, Fang YY, Deng Y, et al. Clinical characteristics of novel coronavirus cases in tertiary hospitals in Hubei Province. Chin Med J (Engl). 2020 Feb 7. doi: 10.1097/CM9.0000000000000744

7. Xu XW, Wu XX, Jiang XG, et al. Clinical findings in a group of patients infected with the 2019 novel coronavirus (SARS-Cov-2) outside of Wuhan, China: retrospective case studies. BMJ. 2020; 368:m606

8. Chan JF, Yuan S, Kok KH, et al. A familial cluster of pneumonia associated with the 2019 novel coronavirus indicating person-to-person transmission: a study of a family cluster. Lancet. 2020; doi: 10.1016/S0140-6736(20)30154-9

9. Gao HN, Lu HZ, Cao B, et al. Clinical findings in 111 cases of influenza A (H7N9) virus infection. N Engl J Med. 2013; 368:2277–85

10. Placzek HED, Madoff LC. Association of age and comorbidity on 2009 influenza A pandemic H1N1-related intensive care unit stay in Massachusetts. Am J Public Health. 2014;104:e118–e125

11. Mauskopf J, Klesse M, Lee S, Herrera-Taracena G. The burden of influenza complications in different high-risk groups. J Med Economics. 2013;16:264–77

12. Shiley KT, Nadolski G, Mickus T, et al. Differences in the epidemiological characteristics and clinical outcomes of pandemic (H1N1) 2009 influenza, compared with seasonal influenza. Infect Control Hosp Epidemiol. 2010; 31: 676–682

13. Martinez A, Soldevila N, Romeo-Tamarit A, et al. Risk factors associated with severe outcomes in adult hospitalized patients according to influenza type and subtype. Plos One. 2019;14:e0210353

14. Gutiérrez-González E, Cantero-Escribano JM, Redondo-Bravo L, et al. Effect of vaccination, comorbidities and age on mortality and severe disease associated with influenza during the season 2016–2017 in a Spanish tertiary hospital. J Infect Public Health. 2019;12:486–491

15. Booth CM, Matukas LM, Tomlinson GA, et al. Clinical features and short-term outcomes of 144 patients with SARS in the greater Toronto area. JAMA. 2003;289:2801–2809

16. Alqahtani FY, Aleanizy FS, Ali Hadi Mohammed R, et al. Prevalence of comorbidities in cases of Middle East respiratory syndrome coronavirus: a retrospective study. Epidemiol Infect. 2018;5:1–5

17. Badawi A, Ryoo SG. Prevalence of comorbidities in the Middle East respiratory syndrome coronavirus (MERS-CoV). Int J Infect Dis. 2016;49:129–133

18. Rahman A, Sarkar A. Risk Factors for Fatal Middle East Respiratory Syndrome Coronavirus Infections in Saudi Arabia: Analysis of the WHO Line List, 2013-2018. Am J Public Health. 2019;305186

19. Alanazi KH, Abedi GR, Midgley CM, et al. Diabetes Mellitus, Hypertension, and Death among 32 Patients with MERS-CoV Infection, Saudi Arabia. Emerging Infect Dis. 2020;26:166–168

20. Yang YM, Hsu CY, Lai CC, et al. Impact of Comorbidity on Fatality Rate of Patients with Middle East Respiratory Syndrome. Sci Rep. 2017;7:11307

21. Garbati MA, Fagbo SF, Fang VJ, et al. A Comparative Study of Clinical Presentation and Risk Factors for Adverse Outcome in Patients Hospitalised with Acute Respiratory Disease Due to MERS Coronavirus or Other Causes. Plos One. 2016;11:e0165978

22. Rivers CM, Majumder MS, Lofgren ET. Risks of Death and Severe Disease in Patients With Middle East Respiratory Syndrome Coronavirus, 2012–2015. Am J Epidemiol. 2016;184:460–464

23. Kulscar KA, Coleman CM, Beck S, Frieman MB. Comorbid diabetes results in immune dysregulation and enhanced disease severity following MERS-CoV infection. JCI Insight. 2019;20:e131774

24. Matsuyama R, Nishiura H, Kutsuna S, et al. Clinical determinants of the severity of Middle East respiratory syndrome (MERS): a systematic review and meta-analysis. BMC Public Health. 2016;16:1203

25. WHO. Clinical management of severe acute respiratory infection when Novel coronavirus (nCoV) infection is suspected: interim guidance. Jan 28, 2020. https://www.who.int/internal-publications-detail/clinical-management-of-severe-acute-respiratory-infection-when-novel-coronavirus-(ncov)-infection-is-suspected (accessed February 25th, 2020)

26. Metlay JP, Waterer GW, Long AC, et al. Diagnosis and treatment of adults with community-acquired pneumonia: An official clinical practice guideline of the American Thoracic Society and Infectious Disease Society of America. Am J Respir Crit Care Med. 2019; 200:e45–e67

27. Fang L, Gao P, Bao H, et al. Chronic obstructive pulmonary disease in China: a nationwide prevalence study. Lancet Respir Med. 2018;6:421–430

28. Moni MA, Lionel P. Network-based analysis of comorbidities risk during an infection: SARS and HIV case studies. BMC Bioinformatics 2014, 15:333

29. Naqvi AA, Shah A, Ahmad R, Ahmad N. Developing an Integrated Treatment Pathway for a Post-Coronary Artery Bypass Grating (CABG) Geriatric Patient with Comorbid Hypertension and Type 1 Diabetes Mellitus for Treating Acute Hypoglycemia and Electrolyte Imbalance. J Pharm Bioallied Sci. 2017;9:216–220

30. Murphy TE, McAvay GJ, Allore HG, et al. Contributions of COPD, asthma, and ten comorbid conditions to health care utilization and patient-centered outcomes among US adults with obstructive airway disease. Int J Chron Obstruct Pulmon Dis. 2017;12:2515–2522

